## Supplementary figures and images for "The Oxford Brain Health Clinic: Protocol and Research Database"

### Figure 1 Greyscale

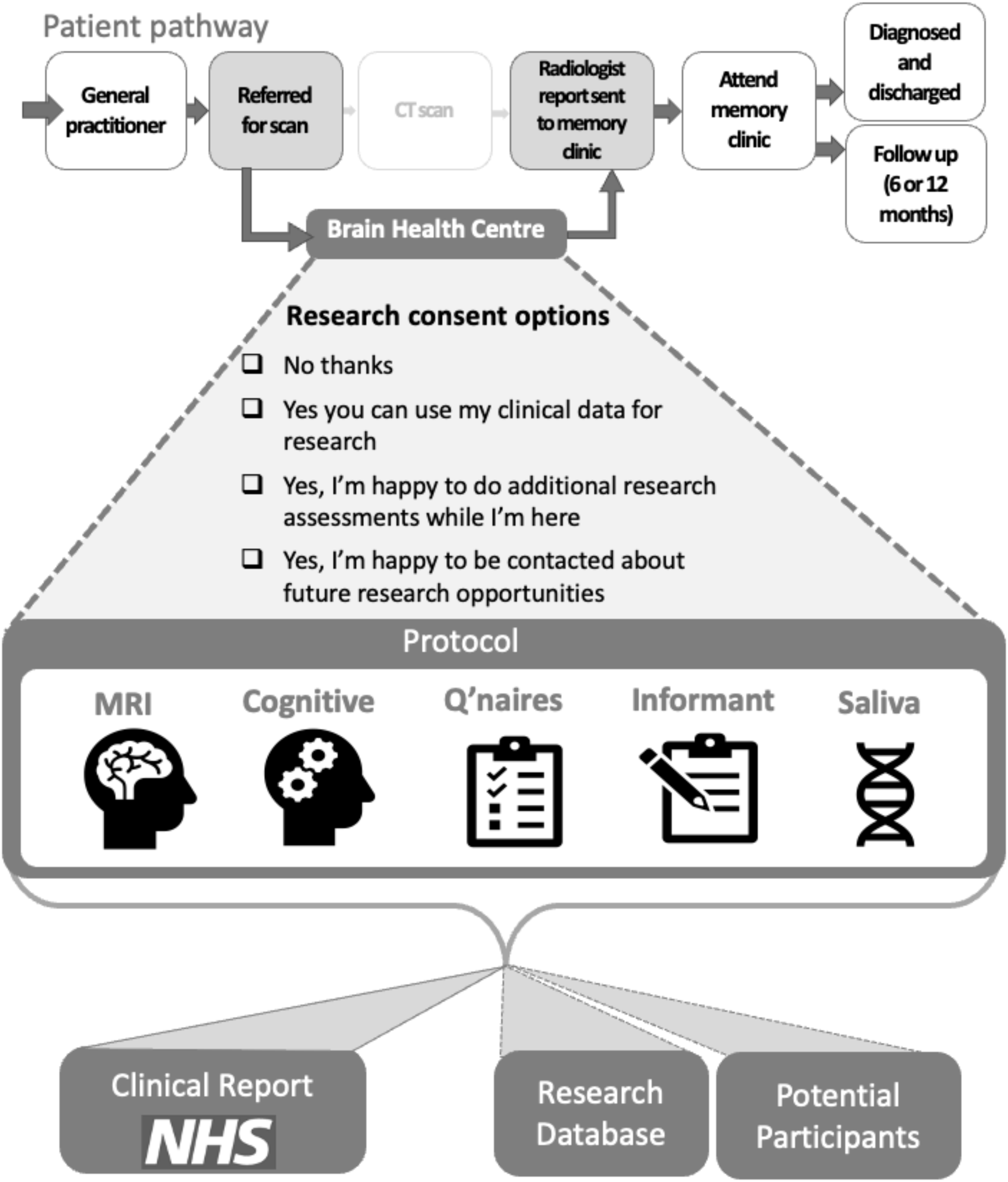

### Figure 2 Greyscale

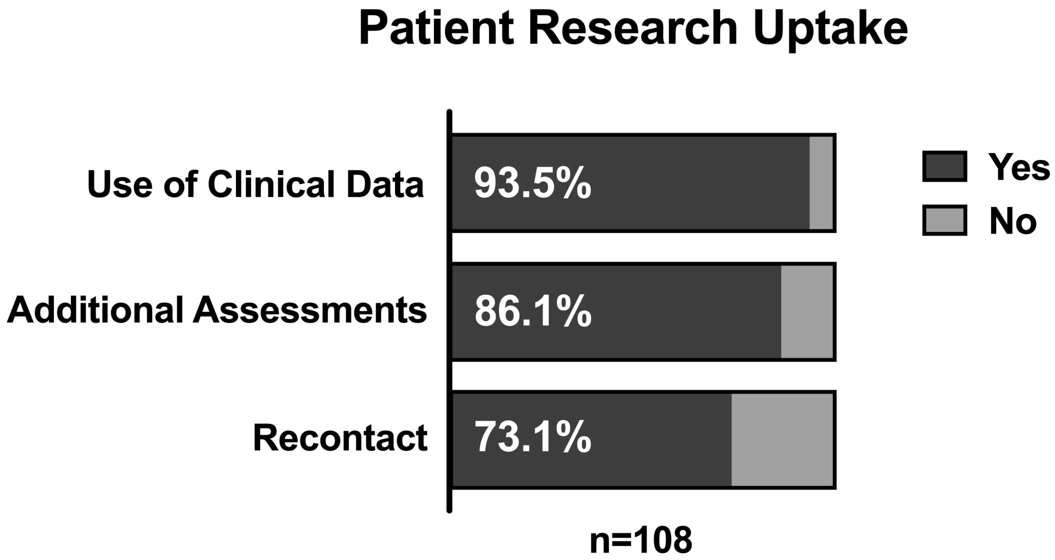

### Figure 3 Greyscale

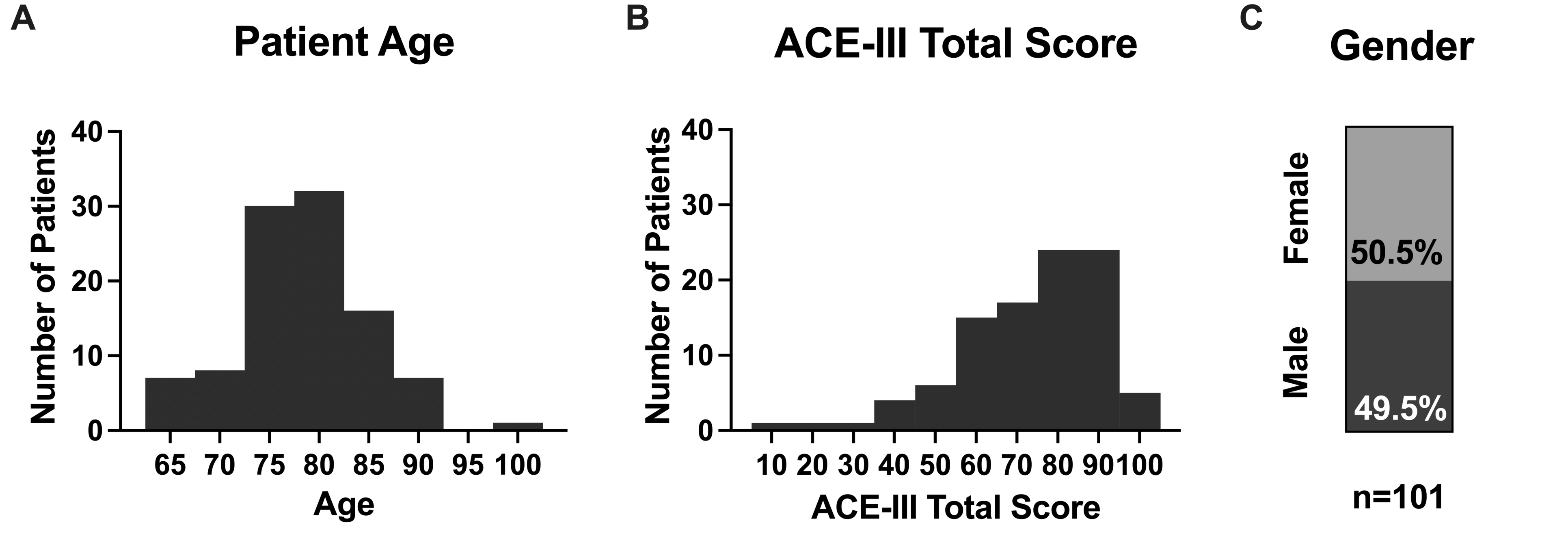

### Figure 4 Greyscale

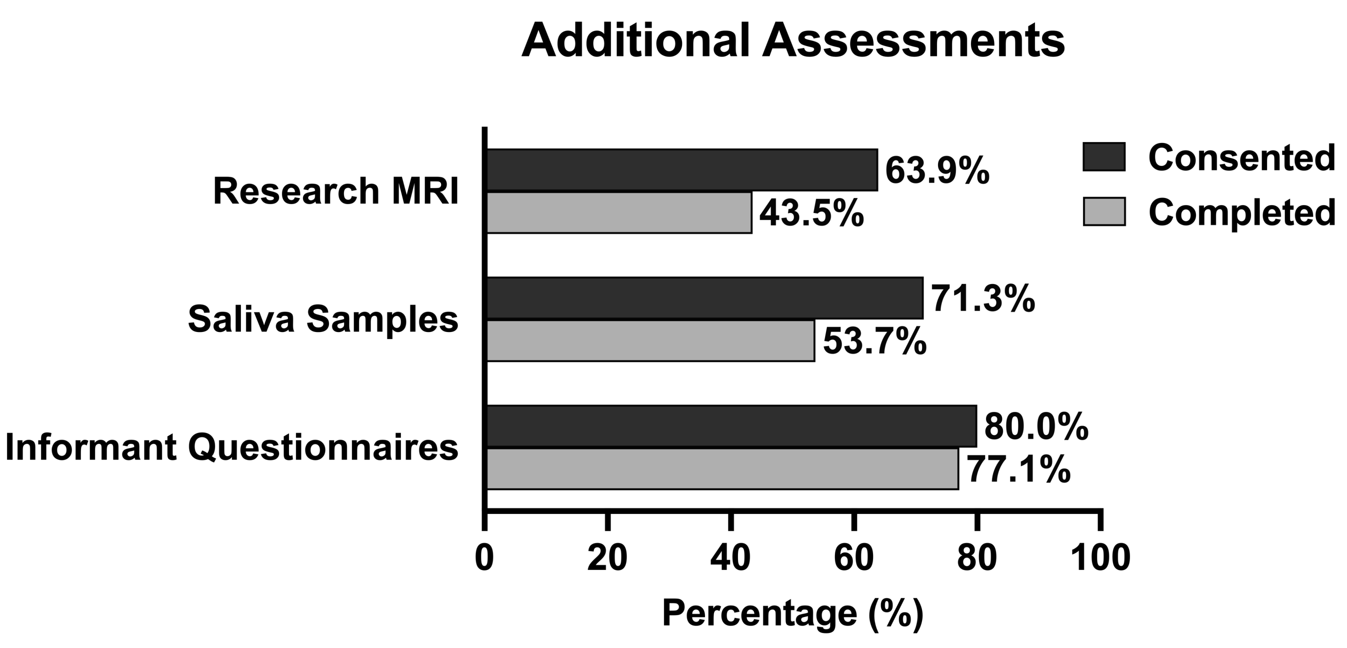
